# A single center cross-sectional study on the efficacy of low-dose cytarabine and aclramycin combined with granulocyte colony-stimulating factor (CAG regimen) in elder adults with acute myeloid leukemia

**DOI:** 10.1101/2024.11.18.24317527

**Authors:** MinXu, Chunxiao He, Xuemei Wen, Chen Lan, Xintong Li, Yaqun Hong, Xiaofan Li

## Abstract

**Background:** Nowadays, the combination of CAG regimen with targeted therapy and immunotherapy has greatly improved the prognosis of AML patients, but there are controversies about the prognostic factors of CAG regimen alone, especially in AML elders.

**Objective:** By investigating the survival status of elder patients who received induction CAG regimen in our hospital at the beginning of this century and analyzing the factors affecting survival, we aim to provide scientific evidence for improving the survival of current patients.

**Method:** The AML elder patients treated with induction CAG regimen including low-dose cytarabine (10 mg/m^2^ per 12 hours, day 1 to 14), aclarubicin(14 mg/m ^2^ per day, day 1 to 4), and G-CSF priming (200 ug / m^2^ per day, day 1 to 14) in Fujian Medical University Union Hospital from January 2001 to December 2009 were involved in this research.

**Results:** Among 92 elderly AML patients, 44 (47.8%) showed clinical efficacy, while 48 (52.2%) experienced treatment failure (including 12 deaths (13.0%)). The main adverse reactions of chemotherapy were bone marrow suppression, with mild non hematological adverse reactions. The median recurrence time was 7 months. The risk factors related efficacy were high blasts, elevated LDH and HBDH.

**Conclusion:** The CAG regimen is suitable for elderly patients and can be used for the treatment of relapsed refractory AML and secondary AML. The CAG regimen has mild non hematological adverse reactions. After chemotherapy, there is a longer bone marrow suppression period and a higher infection rate; The reason for the improved efficacy of CAG regimen in current patients may be related to the effective reduction of tumor burden during induction combined with target or immunotherapy.

## Introduction

Acute myeloid leukemia (AML) is a common hematological malignancy that endangers human life. In recent years, there has been progress in the treatment of AML, and new treatment methods such as hematopoietic stem cell transplantation, immunotherapy, and cell therapy have brought new hope to AML. The prognosis of acute myeloid leukemia (AML) has significantly improved, with a complete remission rate of 70% -85% in newly diagnosed adult AML. But the prognosis of elderly AML is still poor. Chemotherapy drugs not only kill leukemia cells, but also cause toxic damage to normal cells and tissues. Conventional or high-dose chemotherapy can cause severe bone marrow suppression and damage to important organs such as the heart, liver, and kidneys in AML patients, especially in the elderly.

The CAG regimen combines granulocyte colony-stimulating factor (G-CSF), low-dose cytarabine (Ara-C), and acaramycin (Acla). Due to its unique pharmacological properties, the combination of G-CSF pre stimulation with Ara-C and Acla can not only drive the G0 phase AML cells into the proliferation cycle, but also increase intracellular AraC metabolism and Ara CTP levels, thereby increasing anti leukemia efficacy. The dosage is small and almost non hematological toxicity, making it suitable not only for the treatment of refractory relapsed and secondary AML, but also for the treatment of elderly patients. Nowadays, the combination of CAG regimen with targeted therapy and immunotherapy has greatly improved the prognosis of AML patients, but there are controversies about the prognostic factors of CAG regimen alone, especially in AML elders.

In this study, we curated a single center cross-sectional research of the survival status of elder patients who received induction CAG regimen in our hospital at the beginning of this century. We analyze the case composition, treatment efficacy, adverse reactions, and high-risk factors of CAG chemotherapy regimen, and follow up patients who achieve complete remission (CR). We aim to find the factors affecting survival and provide scientific evidence for improving the survival of current patients.

## Method

### Study participants and diagnostic criteria

#### (1) Participants

The AML elder patients (age≥60) treated with induction CAG regimen including low-dose cytarabine (10 mg/m^2^ per 12 hours, day 1 to 14), aclarubicin(14 mg/m ^2^ per day, day 1 to 4), and G-CSF priming (200 ug / m^2^ per day, day 1 to 14) in Fujian Medical University Union Hospital from January 2001 to December 2009 were involved in this research. The G-CSF was started 2 hours before the first injection of Ara-C and stopped 12 hours before the last injection of Ara-C (1. When the white blood cell count is greater than 10 × 10^9^/L, the dose of G-CSF is halved; 2.When WBC>20 × 10^9^/L, pause the use of G-CSF and wait for the white blood cells to fall back; 3. If peripheral white blood cells remain low even after discontinuing Ara-C, continue using G-CSF until granulocytes return to normal.)

#### (2) Inclusion criteria

Patients who meet the diagnostic criteria for acute myeloid leukemia and are treated with the induction CAG regimen.

#### (3) Diagnostic criteria

1. All patients were diagnosed according to the clinical manifestations, blood tests, and bone marrow tests in the “Diagnosis and Treatment Criteria for Hematological Diseases”^[1]^.
2. The evaluation of the general condition of patients is based on the scoring criteria of the Eastern Cooperative Oncology Group (ECOG) in the United States^[2]^.
3. Relapsed AML : patients with blasts in the bone marrow exceeding 5%.
4. Refractory AML as described in our previous publication.

#### (4) Exclusion criteria

1. Without completing the chemotherapy regimen
2. Using CAG regimen for consolidation

#### (5) Supportive care

The patient stays in a regular ward and undergoes air ultraviolet disinfection twice a day; Actively control basic diseases (including actively control hypertension and diabetes); Supportive care with antibiotics and transfusions is considered the standard of care.

All extracted data were deidentified in accordance with ethical standards. This study was reviewed by Fujian Medical University Union Hospital Ethical Review Board.

### Efficacy assessment

According to Zhang Zhinan’s “Diagnosis and Efficacy Standards for Hematological Diseases”, the efficacy is divided into complete remission (CR), partial remission (PR), and non remission (NR). The sum of CR rate and PR rate is the total effective rate.

### Adverse reaction determination

Evaluate chemotherapy adverse reactions using the WHO grading system for adverse reactions.

### Statistical analysis

SPSS13.0 statistical software was used for statistics. The analysis of variance of multivariate data is used to compare the WBC, HB, PLT, peripheral blasts and ALB, LDH, HBDH data. Cox regression was used to screen prognostic factors.

## Results

### Patients’ characteristics

A total of 92 patients were in this study, including 54 males (58.7%) and 38 females (41.3%). The average age is (65.36 ±2.55) years. Among them, 78 cases started with anemia symptoms. The patients often have multiple underlying diseases, including 8 cases of chronic bronchitis and emphysema, 8 cases of pulmonary tuberculosis (2 are active), and 2 case of bronchiectasis; Those complicated with cardiovascular diseases include (20 cases of hypertension, 8 cases of coronary atherosclerotic heart disease, 4 cases of atrial fibrillation; The patients with combined digestive system diseases included 2 case of hepatitis cirrhosis, 18 cases of chronic cholecystitis; There were 8 cases with type 2 diabetes; There were 4 cases with other tumors (2 case colon cancer, 2 case breast cancer). Among those patients, 20 had infections in different locations before chemotherapy. According to diagnostic criteria, there were 18 cases of relapse AML (19.6%), 60 cases of refractory AML (65.2%). Sixteen cases (17.4%) were transformed from myelodysplastic syndrome (MDS).

### Efficacy

Fourty-four cases (47.8%) were clinically effective (including 28.3% of CRs and 19.6% of PRs). Among the 26 CR patients, there were 18 cases of refractory AML,8 cases of relapse AML. Those 12 patients who died after treatment all developed severe infections, including 6 cases developed fatal hemorrhage.

### Adverse effect

The main non hematological adverse effect were nausea, vomiting, decreased appetite (15.2%), fatigue (28.3%), muscle soreness (37.0%), and oral ulcers (8.7%). The above symptoms can be tolerated by patients, and after symptomatic treatment, the symptoms and signs can disappear in a short period of time. Four cases of mild reversible liver dysfunction occurred after chemotherapy. None of the patients experienced renal dysfunction or neurotoxic reactions.

All patients experienced a period of bone marrow suppression(10.38 ± 5.33 days). The lowest WBC,PLT counts and hemoglobin were 0.90 ± 0.53(× 10^9)^, 15.70 ± 10.18(× 10^9)^ and 61.55 ± 9.49(g/L)respectively. During the bone marrow suppression period, the average RBC transfusion volume was 7.58 ± 5.08 units, and the average platelet transfusion volume was 25.0 ± 27.3 units. Sixty-eight patients developed fever during or after chemotherapy (including 40 patients with fever ≤ 39 °C and 28 patients with fever>39 °C), with a median duration of 8.5 days (4-20 days). The infection in respiratory tract, intestinal tract, urinary tract and bloodstream were 78.3%,19.6% 4.3% and 2.2% respectively. Bleeding may occur due to low platelets, including skin bleeding (30.4%), nasal bleeding (6.5%), intracranial bleeding (8.7%), conjunctival bleeding (6.5%), gingival bleeding (2.2%), hemoptysis (4.3%).

### Risk factor

The risk factors related efficacy were high blasts, elevated LDH and HBDH (Tab1). The median recurrence time was 7 months.(Fig1)

**Figure 1.**
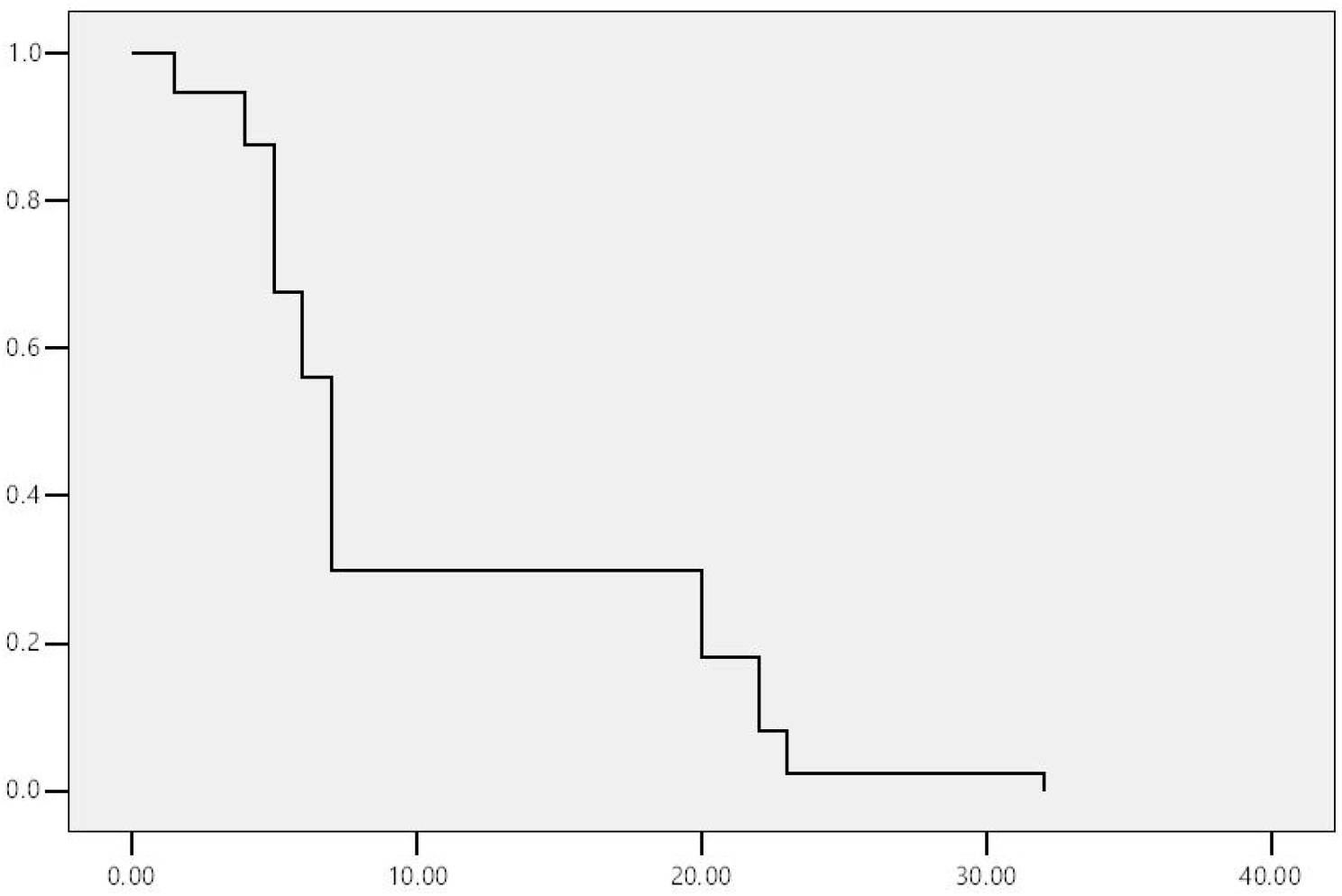
The median recurrence time.

**Table 1.** The risk factors related efficacy.

|  | effective | NR |
| --- | --- | --- |
| Blast(%)* | $17.10 \pm 10.90$ | $41.77 \pm 22.68$ |
| WBC($\times 10^9$) | $6.46 \pm 1.47$ | $5.97 \pm 2.54$ |
| Hb (g/l) | $80.32 \pm 22.02$ | $76.52 \pm 18.42$ |
| PLT ($\times 10^9$) | $74.10 \pm 26.30$ | $55.7 \pm 37.84$ |
| LDH* | $217.24 \pm 91.52$ | $405.19 \pm 108.62$ |
| HBDH* | 202.5 ± 101.26 | 367.23 ± 136.71 |
| albumin | 34.03 ± 5.85 | 34.4 ± 4.71 |

\*P ≤ 0.05

## Discussion

In 1995, Yamada et al. ^[3]^ and Saito et al. ^[4]^ used the proposed CAG regimen to treat patients with relapsed, refractory, and secondary AML, achieving CR rates of 83% and 62%, respectively, with minimal toxic side effects. The above CAG regimen consists of three drugs: Ara-c, Acla, and G-CSF. Ara-c is a S-phase specific drug. After entering the cell, it is converted into its active form, Ara-c triphosphate, by cytosine/deoxyuridine kinase, which interferes with DNA replication and synthesis in S-phase cells. Its metabolite, uridine, can synchronize cells in the S-phase. Acla is a second-generation anthracycline antitumor antibiotic with lipophilic properties, which easily enters cells and maintains high concentrations. It can rapidly transport into the cell nucleus, where it can insert into the DNA double helix, leading to changes in DNA physicochemical properties, inhibiting nucleic acid synthesis (especially RNA synthesis), and arresting cells in the G1 and late S phases^[5]^. G-CSF can specifically induce the proliferation and differentiation of granulocyte progenitor cells, and has a significant effect on promoting the formation of colonies of granulocyte-committed cells (CD34 + /CD33+) in the bone marrow, with a phase of action in the late stage of hematopoietic cell differentiation. G-CSF can significantly increase the colony formation of AML cells, improve the DNA synthesis of AML cells, promote the leukemia cells to enter the S phase, and increase the efficacy of S-phase specific chemotherapy drugs ^[6]^. In addition, G-CSF can accelerate the recovery of neutrophils after chemotherapy, shorten the granulocytopenia period after chemotherapy, and reduce the occurrence of severe infections.

The combination of three drugs in the CAG regimen not only leverages the pharmacological properties of each drug, but also forms a rational combination in terms of drug cell kinetics. Research findings indicate that the combination of CAG drugs may have mechanisms of inducing apoptosis, inducing tumor cell differentiation, and downregulating the BCL-2 gene ^[7-9]^. Chemotherapy drugs have poor killing effect on cells in the G0 phase, while G-CSF can promote G0 leukemia cells to enter the cell proliferation cycle, thereby enhancing the cytotoxic effect of cycle-specific drugs^[10]^.Bai et al. found that the CAG regimen played an important role in inducing apoptosis during the eradication of leukemia cells, and Acla was effective against multidrug-resistant (MDR) cells, with the presence of G-CSF enhancing its effect ^[11]^.

Since Yamada reported the CAG regimen, Japan has conducted several small-scale clinical trials for the treatment of refractory AML, with CR rates of 62%-86% and median survival times of 8-17 months. Compared with conventional chemotherapy, it has fewer toxic side effects, with only mild to moderate bone marrow suppression, and high quality of life for patients; The treatment effect of relapsed AML is particularly good, with a CR rate of 86% and a median survival time of 17 months. Relapsed patients who have undergone CAG treatment after CR can still achieve remission without drug resistance; The efficacy of AML in the elderly was second, with a CR rate of 62.5% and a median survival time of 11 months; The CR rate of secondary AML was 44.4%, with a median survival time of 17 months; The efficacy of primary resistant AML was the worst, with only 1 CR among the 8 patients treated. ^[4]^ The efficacy rate in this article is lower than that reported, which may be related to the following characteristics of this group of patients: ① Most of the patients are elderly, and there are reports that the CR rate of elderly AML patients is inversely proportional to age. If the patient is aged 60-70 years, the CR rate can reach about 50%, and if the patient is aged 70-75 years, the CR rate is about 35%. If the patient is over 75 years old, the CR rate is even lower. ② Multiple underlying diseases are present. ③ Nearly half of the patients had infections in different parts before chemotherapy, and chemotherapy was carried out in parallel with anti-infection treatment, and a large proportion of patients with refractory AML. ④ Most patients have poor general conditions, often accompanied by fever, anemia, low peripheral white blood cell and/or platelet counts before treatment, and have poor tolerance to combined chemotherapy.

In China, there are varying reports on the efficacy of applying the CAG regimen to treat refractory/recurrent AML, with CR rates ranging from 40-75% and effective rates ranging from 60-93%. Shanghai Ruijin Hospital applied the CAG regimen to treat relapsed refractory AML, and the results showed a CR rate of 48.4% (30/62). The 2002 interim summary of the CAG clinical treatment collaboration group in East China included a total of 101 cases of refractory and relapsed AML. The CR rate was 39.6%, the PR rate was 31.2%, and the overall response rate was 71.3%. This article uses the CAG regimen for treatment, with a CR rate of 30.0% and an effective rate of 53.3% in the refractory AML group; The CR rate of the relapsed AML group was 44.4%, and the effective rate was 55.6%, which was slightly lower than the literature reports, which may be related to the case composition. At present, refractory AML is still a difficult point in leukemia treatment. There are different reports on the CR rate of refractory/recurrent AML treatment schemes at home and abroad. The CR rate of IDA scheme in the first-line scheme can reach 45-80% ^[12,13]^, and the CR rate of medium and high dose cytarabine combined with second-line chemotherapy drugs can reach about 40-60% ^[14,15]^. The CR rate of ME and MEA schemes is 30-53%, and the effective rate is about 50-70% ^[16,17]^. The CAG regimen has good efficacy in the treatment of refractory AML and can be used for patients who cannot tolerate medium and high-dose chemotherapy, especially elderly patients.

In terms of adverse reactions, most reports indicate that the CAG regimen has mild side effects. This article shows that non-hematological adverse reactions are very mild and are safer than standard regimens. Despite this, there are still a few reports of serious adverse events induced by toxic side effects: low-dose Ara-c combined with G-CSF can cause severe bone marrow suppression,such as a longer period of granulocytopenia; Taguchi et al. ^[18]^reported two cases of aplastic anemia transformed into AML. After treatment with the CAG regimen, tumor cells significantly decreased, but persistent pancytopenia persisted, resulting in one death from pneumonia and one death from central bleeding. On the other hand, there are reports that low-dose Ara-c combined with G-CSF can cause skin infiltration and an increase in primitive cells in peripheral blood. This article shows that the CAG regimen chemotherapy has a longer period of hematological bone marrow suppression and a higher infection rate after chemotherapy, which supports the importance of supportive therapy after chemotherapy. All these indicate that the clinical application of CAG regimen still needs to be cautious, and cases should be selected carefully.

In terms of risk factors, it is generally believed that factors such as LDH, HBDH have a significant impact. In the study, it was found that there were statistically significant differences in the high blasts, elevated LDH and HBDH between the different efficacy (effective and NR) groups.

Currently, the combination of CAG regimen with targeted therapy and immunotherapy has significantly improved the prognosis of AML patients. The cross-sectional data from this study holds great significance for current treatments. Tumor burden plays a crucial role in treatment efficacy, and the integration of targeted therapy, immunotherapy, and CAG regimen can reduce tumor burden, thereby prolonging the survival time of patients.For CAG regimen combined with immunotherapy and targeted therapy, recent studies have shown that decitabine combined with CAG regimen has a definite effect on the treatment of elderly AML, which can effectively reduce serum VEGF, TSGF, TGF-β1 levels, improve immune function, effectively inhibit the formation of new blood vessels, and prolong the survival of patients^[19,20]^.Decitabine combined with modified CAG regimen is safe and effective in the treatment of AML patients ≥70 years old, and can partially improve the prognosis of elderly and high-risk patients^[21]^.Yifan Liu et al. ^[22]^showed that the VEN-DCAG regimen may be an effective treatment option for patients with R/R AML, with higher rates of ORR and MRD-negative remission in both the relapsed and refractory groups. One limitation of our study stemmed from the inadequately sized dataset. it is imperative to expand the dataset’s scale. This expansion will bolster the resilience of COX models on a constrained dataset from a single center. The other one is due to the cross-sectional study, the MICM was not widely applied at that time.

## Conclusions

The CAG regimen is suitable for elderly patients and can be used for the treatment of relapsed refractory AML and secondary AML. The CAG regimen has mild non hematological adverse reactions. After chemotherapy, there is a longer bone marrow suppression period and a higher infection rate; The reason for the improved efficacy of CAG regimen in current patients may be related to the effective reduction of tumor burden during induction combined with target or immunotherapy.

## Data Availability

All data produced in the present study are available upon reasonable request to the authors

## Acknowledgment

The authors thank the patients and their families, all the investigators and the staffs involved in data collection and analyses.

## Disclosure of conflict of interest

All authors agree that there is no conflict of interest in publishing this manuscript.

## Authorship contribution

Conception and design: Xiaofan Li.

Provision of study materials and patients:MinXu and Chunxiao He.

Collection and assembly of data: All authors.

Data analysis and interpretation: MinXu,Chen Lan and Yaqun Hong.

Manuscript writing: Chunxiao He and Xuemei Wen.

Approval of the manuscript: All authors.

Accountable for all aspects of the work: All authors.

## Data and code availability

The main data supporting this study is included in the article/supplementary material. Further inquiries can be directed to the corresponding authors.

## Funding

This work was supported by grants from Fujian Province Youth Top Talent Project; National Key R&D Program of China[2022YFC2502704];Fujian Provincial Health Commission Young and Middle-aged Talent Project[2021GGA018];Natural Science Foundation of Fujian Province funding project[2021J01782]to XiaofanLi.

## Notes

### Competing Interest Statement

The authors have declared no competing interest.

### Funding Statement

This study was funded by grants from Fujian Province Youth Top Talent Project; National Key R&D Program of China[2022YFC2502704];Fujian Provincial Health Commission Young and Middle-aged Talent Project[2021GGA018];Natural Science Foundation of Fujian Province funding project[2021J01782]to XiaofanLi;

### Author Declarations

by Fujian Medical University Union Hospital Ethical Review Board

